# Performance of Saliva, Oropharyngeal Swabs, and Nasal Swabs for SARS-CoV-2 Molecular Detection: A Systematic Review and Meta-analysis

**DOI:** 10.1101/2020.11.12.20230748

**Authors:** Rose A. Lee, Joshua C. Herigon, Andrea Benedetti, Nira R. Pollock, Claudia M. Denkinger

**Author notes:** Corresponding Author: Nira Pollock, M.D., Ph.D., D(ABMM), Associate Medical Director, Infectious Diseases Diagnostic Laboratory, Boston Children’s Hospital, Division of Infectious Diseases, Beth Israel Deaconess Medical Center, Associate Professor of Pathology and Medicine, Harvard Medical School Farley Building 8th floor, Room FA828, 300 Longwood Ave 45 Boston, MA 02115.

## Abstract

**Background:** Nasopharyngeal (NP) swabs are considered the highest-yield sample for diagnostic testing for respiratory viruses, including SARS-CoV-2. The need to increase capacity for SARS-CoV-2 testing in a variety of settings, combined with shortages of sample collection supplies, have motivated a search for alternative sample types with high sensitivity. We systematically reviewed the literature to understand the performance of alternative sample types compared to NP swabs.

**Methods:** We systematically searched PubMed, Google Scholar, medRxiv, and bioRxiv (last retrieval October 1st, 2020) for comparative studies of alternative specimen types [saliva, oropharyngeal (OP), and nasal (NS) swabs] versus NP swabs for SARS-CoV-2 diagnosis using nucleic acid amplification testing (NAAT). A logistic-normal random-effects meta-analysis was performed to calculate % positive alternative-specimen, % positive NP, and % dual positives overall and in sub-groups. The QUADAS 2 tool was used to assess bias.

**Results:** From 1,253 unique citations, we identified 25 saliva, 11 NS, 6 OP, and 4 OP/NS studies meeting inclusion criteria. Three specimen types captured lower % positives [NS (0.82, 95% CI: 0.73-0.90), OP (0.84, 95% CI: 0.57-1.0), saliva (0.88, 95% CI: 0.81 – 0.93)] than NP swabs, while combined OP/NS matched NP performance (0.97, 95% CI: 0.90-1.0). Absence of RNA extraction (saliva) and utilization of a more sensitive NAAT (NS) substantially decreased alternative-specimen yield.

**Conclusions:** NP swabs remain the gold standard for diagnosis of SARS-CoV-2, although alternative specimens are promising. Much remains unknown about the impact of variations in specimen collection, processing protocols, and population (pediatric vs. adult, late vs. early in disease course) and head-to head studies of sampling strategies are urgently needed.

## Introduction

Testing for SARS-CoV-2 (severe acute respiratory syndrome coronavirus 2, the etiologic agent of COVID-19), has preferentially utilized nasopharyngeal (NP) sampling with flocked swabs. This sampling method is presumed to have the highest diagnostic yield, as evidenced by its use as a reference method by the Food and Drug Administration (FDA).(1). However, NP sampling requires professional collection (a major limitation given the strained health care system in almost all parts of the world) and protective equipment which, similar to the flocked swabs themselves, is in short supply. In addition, NP sampling is uncomfortable, limiting patients’ willingness to come forward for testing especially if asymptomatic.

If large scale testing of symptomatic and asymptomatic patients is to become a reality, innovation in sampling is as important as innovation in testing. Innovation in sampling requires consideration of sampling methods (e.g., choice of swab, choice of sample type) as well as self-sampling.

As of October 2020, interim guidance from the Centers for Disease Control and Prevention (CDC) (2) recommends upper respiratory tract testing with any of the following specimens: NP swab, NP wash/aspirate, nasal wash/aspirate, oropharyngeal (OP) swab, nasal mid-turbinate (MT) swab using a flocked tapered swab, an anterior nares (AN) nasal swab using a flocked or spun polyester swab, or a saliva specimen obtained by supervised self-collection. The FDA, in contrast, states that NP, OP, MT, and AN swab samples are appropriate for clinical testing, and that “more data are necessary to better understand the performance when using specific saliva collection devices or other specimen types for COVID-19 testing” (1).

Saliva is the subject of the largest body of research on alternative specimen types as well as the specimen of choice for numerous companies putting high-volume testing programs in place. Certain regions of the world, such as Hong Kong, have already adopted saliva in their mass screening protocols (3). The pathophysiological rationale for saliva sampling is based on the angiotensin-converting enzyme II (ACE-2) being the cellular receptor for SARS-CoV-2 (4), similar to SARS-CoV (5). High ACE-2 receptor expression in oral mucosa and salivary glands has been recently demonstrated (6, 7). Salivary gland duct epithelial cells previously were identified as a target for SARS-CoV in a rhesus macaque model (8). One study detected SARS-CoV-2 in saliva collected via expression directly from the salivary gland duct (9). These findings suggest that saliva may be a suitable and high-yield diagnostic sample for SARS-CoV-2 testing based on local viral replication, in addition to the possible mixing in saliva of lower and upper respiratory tract fluids that can carry virus. Saliva offers several advantages as it is a non-invasive sample type that can be self-collected, thus decreasing infectious risk to medical personnel, use of personal protective equipment, and reliance on equipment subject to supply shortages such as nasal, OP, or NP swabs.

OP (throat) swabs have been in wide use for SARS-CoV-2 diagnosis since the beginning of the pandemic (3). Consensus is lacking on both the best collection approach and nomenclature for oral specimens. The terms “oropharyngeal” and “throat” have been used in the literature, and therefore we describe and group them here as “oropharyngeal “or OP swabs. No comprehensive evaluations of their performance compared to other methods exist. OP swabs are less specialized than NP swabs and thus OP samples can be collected with a broader range of swab products. While the FDA recommends that OP swabs be collected by a health care professional (1), some have suggested that self-swabbing might be possible. Nasal swabs (NS), another important alternative specimen type, also have the advantages of increased comfort and possible self-collection. They have been classified into two types anatomically. Nasal mid-turbinate (MT) swabs, also called deep nasal swabs, are defined by the CDC (2) as flocked/tapered swabs performed while tilting the patient’s head back 70 degrees and inserting the swab less than one inch (about 2 cm into the nostril) until resistance is met at the turbinates before rotating the swab several times against the nasal wall. Anterior nares (AN) swabs are defined without a head tilt and inserting the entire swab at least 0.5 inch (1cm) inside the nostril (naris) and sampling the membrane by various methods, including rotating the swab in place or around the inside wall of the nostril multiple times, and/or leaving in place for 10 to 15 seconds. The current “lower nasal swab” protocol sanctioned by the FDA specifies swab insertion “until you feel a bit of resistance” and thus matches the MT depth defined by the CDC, though swab type is not specified (10). The CDC and FDA suggest swabbing both nares for sampling.

A large body of literature on the yield of these alternative sample types for nucleic acid amplification testing (NAAT) has been created but a comparative systematic summary of the relative performance of these alternative specimen types, compared to NP swabs, is missing and is needed to inform decision makers. Furthermore, the data for self-collection of these alternative specimens (1, 2) have not been systematically evaluated. Here, we aim to clarify the performance of alternative specimen types for diagnosis of SARS-CoV-2 by systematically reviewing and meta-analyzing the literature on this topic available through October 2020.

## Methods

This systematic review and meta-analysis is reported in accordance with PRISMA guidelines (see Supplementary File for the PRISMA checklist). The protocol for this work was registered in the International Prospective Register of Systematic Reviews (PROSPERO) (identifier: CRD42020214660).

### Search strategy, information sources, and eligibility criteria

We performed a comprehensive search of the following databases (Pubmed/MEDLINE and Google Scholar) as well as the preprint servers medRxiv and bioRxiv to identify relevant studies from January 1st, 2020 until October 1st, 2020. Only English language articles were allowed. An example search strategy is provided in Supplementary Methods. Additional studies were retrieved by screening the reference lists of the included articles and from archives of the reviewers. We excluded papers from the same hospital with overlapping inclusion dates, to avoid including patients more than once and thus minimize bias in the data (11). Cross-sectional, case-control, and cohort studies and randomized controlled trials were included independent of number of specimens tested. Conference proceedings and abstracts were deemed ineligible. Participants of all age groups with presumed SARS-CoV-2 infections, in all settings, were included. We included only papers utilizing NAAT for SARS-CoV-2 detection.

### Data Extraction

Two reviewers assessed all articles (R.L. and J.H.) independently and disagreements were resolved with input of a third investigator (N.R.P. or C.M.D.).

We compared alternative sampling to NP sampling. We extracted only data for positive NAAT results on at least one sample type and only when sampling methods being compared were performed synchronously. If patients were tested serially over time and data could not be extracted for specific timepoints, we excluded the study. If multiple different RT-PCR tests were performed on one sample within one study, we utilized the data from the best-performing assay. If a study contributed data to more than one analysis (e.g., two different alternative sample types in one study, each compared to NP swab), it was considered as two or more datasets. If a combined NP/OP sample was used, and no data was available for NP sampling alone, we included the study, using NP/OP as the comparator. For each specimen type, in addition to data on test performance, we extracted data for factors likely to affect test performance as detailed below. Data on throat or gargle washes were not included in this meta-analysis (12, 13).

Our study retrieval process is depicted in Supplementary Figure 1. Records were organized using a reference manager (Zotero Version 5.0.89, George Mason University).

### Assessment of methodological quality

The Quality Assessment of Diagnostic Accuracy Studies-2 (QUADAS-2) tool, a validated quality assessment tool for diagnostic studies, was used to assess the included studies’ risk of bias (14). The four domains assessed for risk of bias and applicability include: 1) participant selection; 2) index test; 3) reference test; 4) flow and timing.

### Data analysis

Under the assumption that 1) we do not know in advance which specimen type performs best for SARS-CoV-2 detection and 2) false positives are very infrequent for NAATs performed in qualified laboratories, we focused only on individuals positive for at least one sample type and report on agreement between samples rather than measures of sensitivity and specificity. Specifically, for each study, we included all individuals with paired specimens who had at least one positive specimen as the denominator, and calculated 1) % positive Alt [percent of individuals positive by the alternative specimen type (e.g. saliva, NS, or OP swab)], 2) % positive NP [percent of individuals positive by the NP specimen (or NP/OP in a minority of cases)], and 3) % dual positive (percent of individuals positive for both the alternative specimen and NP specimen).

We present results of the systematic review in forest plots. Meta-analyses were only performed when there were at least 4 primary studies and 20 patients per sub-group analysis. The % positive Alt, % positive NP, and % dual positive for pooled, overall, and in sub-groups were estimated using a logistic-normal random-effects model (via the ‘metaprop_one’ command with Stata (15)). We applied the Freeman-Tukey Double Arcsine transformation to stabilize variances and score 95% intervals were computed. Heterogeneity was measured by the inconsistency index (I^2^), which describe the percentage of variation across studies due to heterogeneity rather than chance. To investigate possible contributors to heterogeneity, we present sub-group analyses by (1) site of sampling; (2) swab material (flocked, unflocked); (3) sampling procedure; (4) professional vs self-sampling; (5) specimen processing/NAAT (including dilution, nucleic acid extraction procedure, and assay used); (6) populations (e.g. pediatric, asymptomatic); (7) symptom duration prior to testing. The robustness of the meta-analysis to publication bias was assessed by the symmetry of funnel plots.

All analyses and graphs were performed using Stata 15.1 (StataCorp, Texas) and GraphPad 8.5 (Prism, SanDiego).

## Results

Our search yielded 1,253 unique citations, of which 25 were included in the analyses for saliva, 11 for NS, 6 for OP, and 4 for OP/NS swabs. Reasons for exclusions of studies are laid out in Supplementary Figure 1.

The studies we included in the meta-analysis overall had a moderate-high risk of bias according to QUADAS-2 (Supplementary Table 1,2) with studies on NS having a low to moderate risk (Supplementary Table 3). Many studies did not specify patient selection methodology (random or consecutive). Many studies were also comprised of cohorts of known positives (case-control) that were then re-tested with paired specimens and therefore at a high risk of a selection bias limiting applicability of results to the general screening population.

### Saliva

In total, 25 studies (16–40) met our inclusion criteria to assess saliva as an alternative sample type for SARS-CoV-2 diagnosis. The studies cumulatively included 4,528 paired saliva and NP swab specimens, although two studies used a combined NP/OP swab as a comparator (24, 39). An additional 13 studies (3, 9, 41–51) described a performance estimate for saliva as a specimen type, but these studies were excluded from the meta-analysis as the saliva and NP samples were either not collected synchronously, or multiple paired specimens were taken from the same set of patients and the data could not be extracted for a unique patient/time point.

Overall across the 25 studies, we found that the % positive saliva [0.88, 95% confidence interval (CI) 0.81 – 0.93] was lower than the % positive NP (or NP/OP) although not substantially different [0.94 (95% CI 0.90 – 0.98)]. The % dual positive was noticeably lower than either specimen type alone [0.79, (95% CI 0.71 – 0.86), Figure 1], indicating relatively poor agreement. Considerable heterogeneity was also detected (I^2^ 88.6%). This heterogeneity was likely attributable to the variation in study procedures and patient population between studies, as outlined in Supplementary Table 4. Notably, there were no head-to-head studies for any of the comparisons described below for differences in collection, processing, and populations.

**Figure 1:**
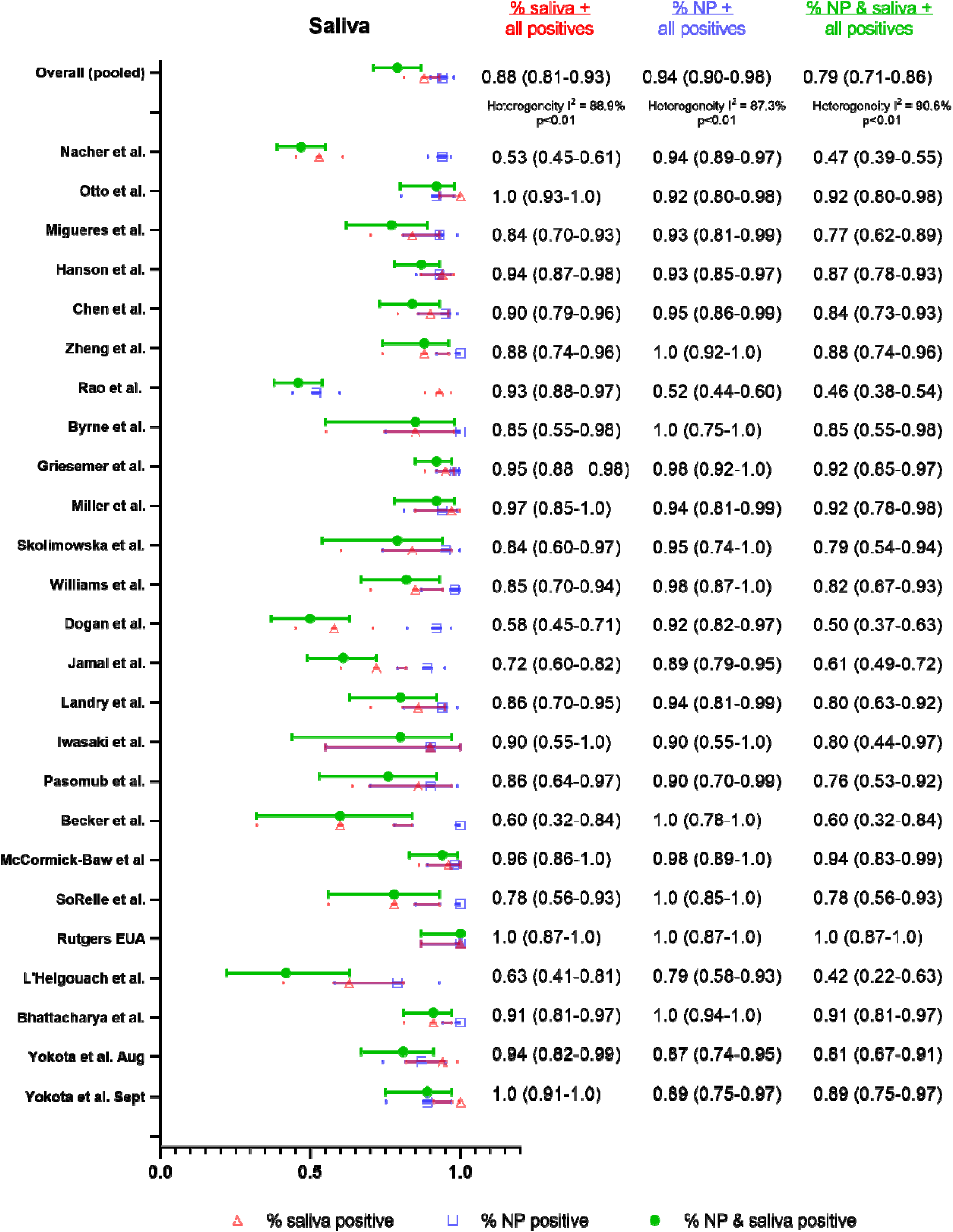
Summary forest plot of individual studies assessing saliva

Saliva collection protocols for included studies were assessed for differences with respect to 1) asking patients to cough or clear their throat before submission of sample (likely mixed sputum and saliva specimen or deep throat saliva specimen) or 2) requesting the patients submit “drool” or “spit”. While some authors (42) have hypothesized that capture of posterior oropharyngeal saliva or mixed sputum/lower respiratory specimen is important for diagnostic sensitivity, we did not find a considerable difference in performance, although % positive saliva was higher for studies (33, 34, 37, 40) that specified cough or deep throat saliva specimen vs studies that did not specifically ask for this [0.94 (95% CI: 0.87-0.99) vs. 0.86 (95% CI: 0.78-0.92), Fig. 2]. For NP samples in these two groups, the % positive detection was similar [0.89 (95% CI: 0.60-1.0) vs. 0.95 (95% CI: 0.93-0.97)]. Notably, many studies that supported the hypothesis of a coughing or deep throat saliva being better than drool/spit sample were excluded from the meta-analysis due to non-synchronous sample collection or repeat testing on the same set of patients (3, 41–43).

**Figure 2:**
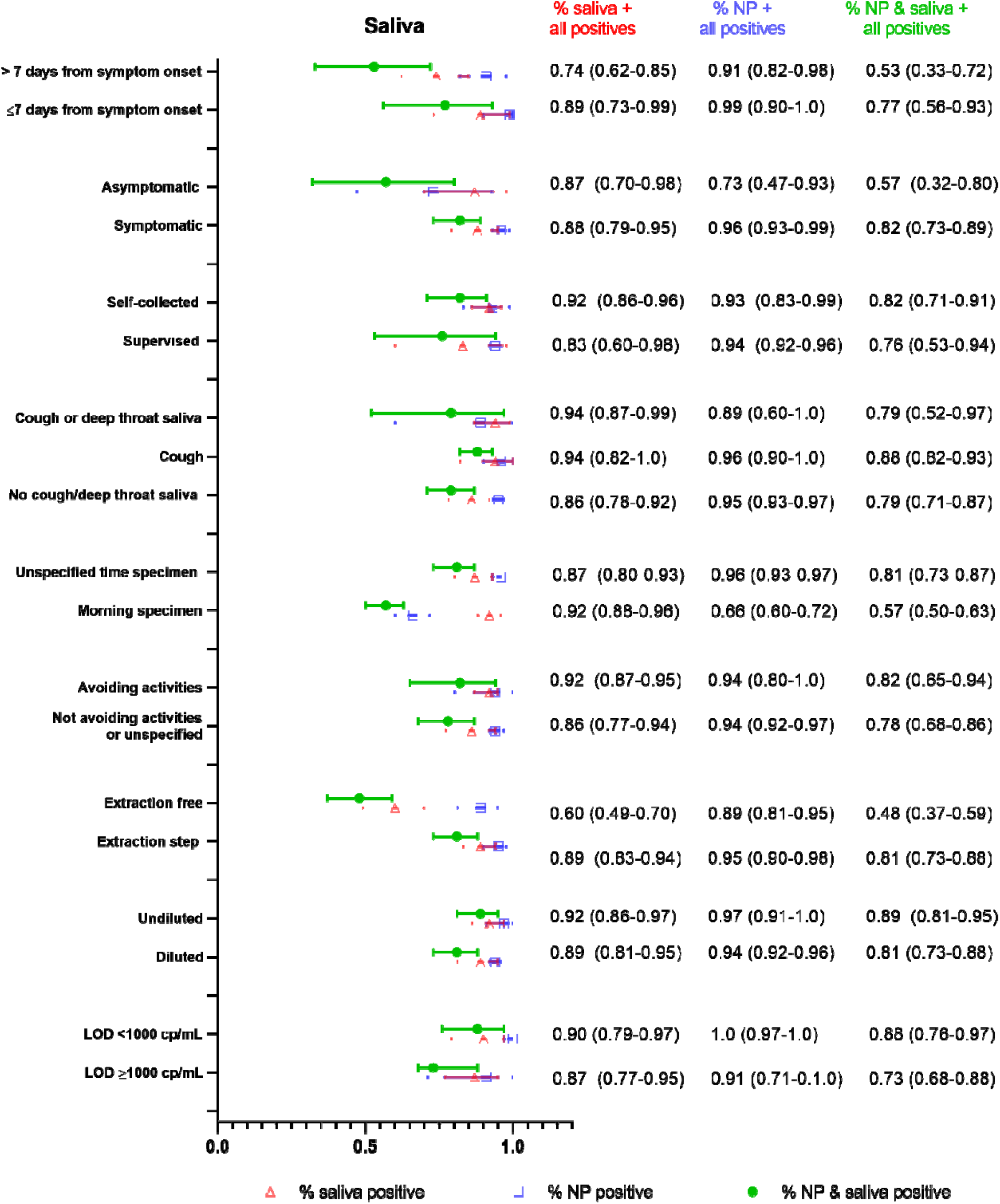
Summary forest plot of sub-group data from saliva sampling for different clinical populations, and collection as well as processing procedures.

Other differences in sampling between studies included avoidance of eating, drinking, or brushing teeth before specimen submission (typically 30 minutes to 2 hours). Protocols that specified avoidance of eating, drinking, or brushing teeth prior to saliva collection (22, 23, 26, 31–34, 40) had higher % positive saliva, although the difference was not substantial [0.91 (95% CI 0.86 – 0.95) vs. 0.86 (95% CI 0.79 – 0.92), Fig. 2]. For NP sampling in the same two groups of studies, the difference was minimal [0.94 (95% CI: 0.8-1.0) vs. 0.95 (95% CI: 0.92-0.97)]. A few studies specifically requested morning submission (33, 40) in addition to avoidance of food/drink and similar saliva performance was identified between protocols that specified morning submission versus those that did not [92% positive saliva (95% CI: 0.88-0.96) vs. 87% (95% CI: 0. 79-0.93), respectively]. However, for NP sampling in the two groups of protocols, the % positive NP was lower with morning submission [66% positive (95% CI: 0.60-0.72) vs. 96% positive (95% CI: 0.94-0.97)], but this was largely driven by one study (33) in which NP swabs performed poorly (this was an outpatient cohort of patients who were asymptomatic at the time of collection and had received their diagnosis at least one week prior).

While many studies specified self-collected saliva (16, 17, 20, 25–27, 29, 30, 33, 34, 37, 39, 40), some studies described supervised collection (22, 28, 31, 35, 38). The % positive saliva was higher for self-collection than supervised collection although the difference was not substantial [0.92 (95% CI: 0.86-0.96) vs. 0.83 (95% CI: 0.60-0.98), respectively, Fig. 2].

There were also substantial differences in specimen processing, including variable dilution of the saliva specimen prior to NAAT, and use of protocols that directly input saliva samples into the NAAT without nucleic acid extraction. We found that even in the absence of dilution for saliva, the % positive could not match that of NP swabs. Studies utilizing undiluted saliva specimens (22, 26) had similar % positive saliva to studies utilizing diluted saliva (16, 17, 23–25, 27–32, 35, 37, 40) [Fig. 2, 0.92 (95% CI: 0.86-0.97) vs. 0.89 (95% CI: 0.81-0.95)]. All NP swabs were eluted in viral transport media, and performance was similar in these two groups of studies [0.97 (95% CI: 0.91-1.0) for undiluted saliva vs. 0.94 (95% CI: 0.92-0.96) for diluted saliva]. In studies using diluted saliva, there was wide variation in dilution methods, with many groups not specifying the degree of dilution.

Studies that did not use a nucleic acid extraction step (19, 28) but instead directly input the saliva specimen into the amplification assay without any pre-processing showed substantially lower % positive saliva than studies that had an extraction step [0.60 (95% CI: 49%-70%) vs. 0.89 (95% CI: 83%-94%), Fig. 2]. In contrast, % positive NP swab was not different between the two groups of studies [0.89 (95% CI: 0.81-0.95) vs. 0.95 (95% CI: 0.90-0.98)], with all except one study (28) using a nucleic acid extraction for the NP swab sample (28).

While the majority of studies utilized reverse transcription polymerase chain reaction (RT-PCR) assays, one study (19) used reverse transcription loop-mediated isothermal amplification (RT-LAMP), and two studies used transcription mediated amplification (TMA) (35, 36). NAATs utilized also differed over ten-fold in their limits of detection between studies, which has been previously described to affect diagnostic positivity (52). We found slightly higher, but not substantially different, percentages of positive detection in studies with a more sensitive/lower [<1000 copies/milliliter (cp/mL)] limit of detection (LOD) (20, 21, 24, 34, 39) compared to studies with LOD ≥ 1000 cp/mL (23, 26, 27, 31–33, 51) [0.90 (95% CI: 0.79-0.97) vs. 0.87 (95% CI: 0.77 – 0.95)]. The differences were similar for NP swabs (100% (95% CI: 0.97-1.0) vs. 0.91 (95%CI: 0.71-1.0)).

We also assessed if patient symptomatology could explain variable diagnostic performance between saliva and NP sampling. We found that only a few studies provided asymptomatic (16, 19, 33)/symptomatic patient data (17, 18, 21–23, 25, 27, 28, 34, 35, 37, 39, 40) that could be parsed and extracted for analysis. We found that % positive saliva was similar between asymptomatic and symptomatic patients [0.87 (95% CI: 0.70-0.98) vs. 0.88 (95% CI: 0.79-0.95) respectively]. The difference between % positive NP in asymptomatic vs symptomatic patients was much larger [symptomatic 0.96 (95% CI: 0.93-0.99) vs. asymptomatic 0.73 (95% CI: 0.47-0.93, Fig. 2)]. These findings, however, were driven by one study (33) with superior saliva performance in asymptomatic patients [% positive saliva 0.93 (95% CI: 0.88-0.97) vs. % positive NP 0.52 (0.44-0.60)]. As discussed above, this study (33) included an outpatient cohort who had received their diagnosis at least one week prior. Notably, although the patient population is described as “asymptomatic,” it is unclear if this was just at the time of collection and they had symptoms closer to their initial diagnosis.

Another important question that we were not able to adequately address from the literature is the performance of saliva in pediatric populations. There are two studies (51, 53) that evaluate the performance of saliva in children, both of which showed worse performance compared to NP swabs [8/11 children positive by NP swab were positive by saliva in one study (51), and in the other study 53% of the children detected by NP swab were also positive by saliva (53)]. However, the timing of saliva collection versus NP swab collection in both studies was unclear, results for asymptomatic vs symptomatic children were not clearly distinguished, and sample processing methods were not clearly described.

Invalid test results were also not consistently reported across studies. Viscosity of saliva was highlighted in some studies as increasing errors in automated pipetting steps, necessitating dilution or biochemical pre-treatment of samples (26, 35, 44).

Given that the fluctuation of viral load over time may differ between saliva and NP samples, and the timepoints patients present themselves for diagnosis may vary, we also assessed differences in diagnostic performance at different timepoints throughout the illness. Sub-group analysis of 6 studies with extractable data (25, 27, 28, 36, 39, 51) found that % positive saliva was overall lower >7 days after symptom onset [0.74 (95%CI: 0.62-0.85)] compared to ≤ 7 days [0.89 (95% CI: 0.73-0.99)], which was also observed for NP swabs [91% (95% CI: 0.82-0.98) vs 99% (95% CI: 0.90-1.0), respectively].

### OP swab

We identified six studies that assessed OP vs NP swabs (54–59) and were suitable for meta-analysis. Given the paucity of data, subgroup analyses to assess differences in collection procedure, sample processing, and patient symptomatology were limited (Supplementary Figure 2). We found that % positive OP swab was similar to % positive NP swab [0.84 (95% CI: 0.57-1.0) vs. 0.88 (95% CI: 0.73-0.98)] although % dual positive was only 0.68 (95% CI: 0.36-0.93) suggesting limited agreement (Figure 3). Notably, the % positive NP estimate was unusually low in this group of studies, which is largely driven by one study (56) with a large gap in performance between % positive OP and % positive NP [0.86 (95% CI: 0.65-0.97) vs. 0.41 (95% CI: 0.21-0.64) respectively] that was not observed in other studies. This study was unique in that paired samples were taken near the end of a hospitalization (unclear duration of symptoms) in a cohort comprised of known positives from prior NP swab RT-PCR positive patients to help determine discharge eligibility.

**Fig. 3:**
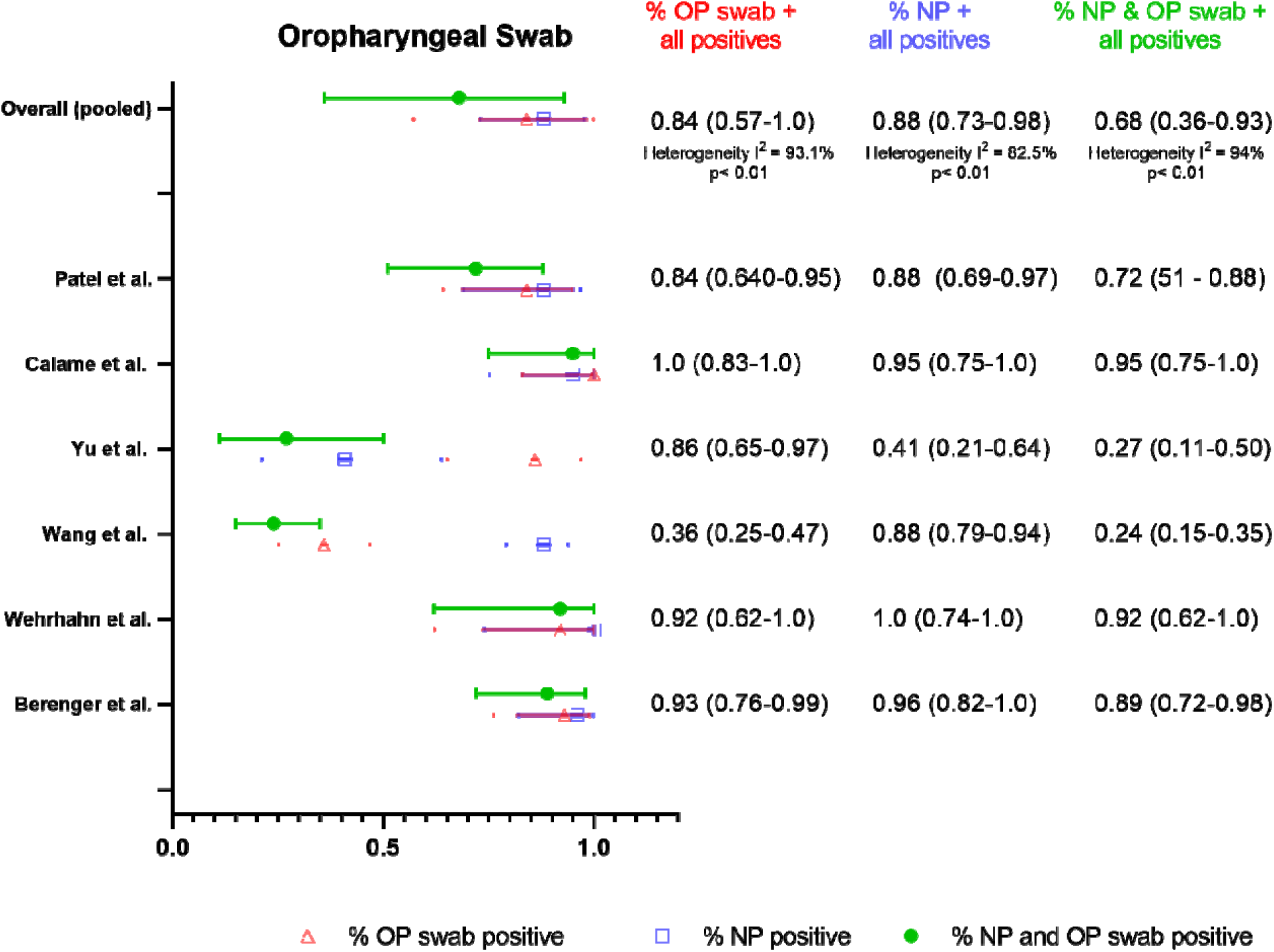
Summary forest plot of individual studies assessing oropharyngeal swabs

Another outlier study in this dataset was a cohort of symptomatic patients (55) in a Wuhan hospital early on in the pandemic with unusually poor performance of OP swabs [0.36 (95% CI: 0.25-0.47)] in comparison to NP swabs [0.88 (95% CI: 0.79-0.94)]. There were not sufficient data in the OP swab meta-analysis to assess the impact of symptomatology or duration of symptoms on percent positives.

Only 3 studies specified flocking of swabs (54, 57, 59), and in all 3 cases unflocked oropharyngeal swabs were compared to flocked NP swabs. The percent positive detection rate for these 3 studies was similar between OP and NP sampling [0.96 (95% CI: 0.88-1.0) vs. 0.97 (95% CI: 0.90-1.0)].

There were two studies that used healthcare-worker collected oral swabs (54, 57) and the positive detection rate for OP vs. NP sampling was similar [0.97 (95% CI: 0.89 – 1.0) vs 0.96 (95% CI: 0.87-1.0) respectively, Fig. 3]. One study used unobserved self-collected OP swabs and reported similar performance between sample types but in a limited sample set of only 12 positive patients [0.92 (95% CI: 0.62-1.0) for OP vs. 1.0 (95% CI: 0.74-1.0) for NP)]. The 3 other studies (55, 56, 58) in this meta-analysis did not specify self-vs. healthcare-worker collection. There was one study (60) that compared self-versus lab-technician collected OP swabs and found that only 14/24 total positives were detected by self-collection versus 22/24 total positives for lab-technician-collected. This study was excluded from this meta-analysis, however, as the pairing of OP swabs to NP swabs was unclear.

Three studies that assessed oral swab specimens were excluded from this meta-analysis as they were not oropharyngeal. There was one pediatric study of buccal swabs in Singapore (61) where children underwent daily NP and buccal swabs; 9/11 children with SARS-CoV-2 detected by NP swab also at some point had positive buccal swabs. One study that described sampling of the anterior 2/3^rd^ of the dorsum of the tongue (62) found similar positive detection rates from “tongue” swabs (46/51 total positives) and NP swabs (49/51 total positives). Another study asked patients to cough prior to sampling oral fluid in cheeks, gums, hard palate, and tongue (63), and reported that detection was slightly higher (26/29 total positives) than NP swabs (23/29 total positives).

### AN and MT Swabs

We identified 11 studies using either AN or MT swab that could be combined for pooled meta-analysis (32, 35, 54, 59, 62–68) with NP swabs as the reference sample type, although one study (59) used a combined NP/OP swab for reference. We found that the % positive NS (either AN or MT) (0.82, 95% CI: 0.73– 0.90), was substantially lower than % positive NP swab (0.98, 95% CI: 0.96 – 1.0) as well as % dual positive (0.79, 95% CI: 0.69 – 0.88), suggestive of limited agreement between the two sample types (Fig. 4). Considerable heterogeneity was again detected between studies (I^2^ = 87%), which we attributed to differences in study procedures. Accordingly, we assessed performance by collection protocol, self-collection/supervision/healthcare-worker-collection, sample processing, and patient symptoms (Supplementary Table 5). Notably there were no head-to-head studies for any of the comparisons.

**Fig. 4:**
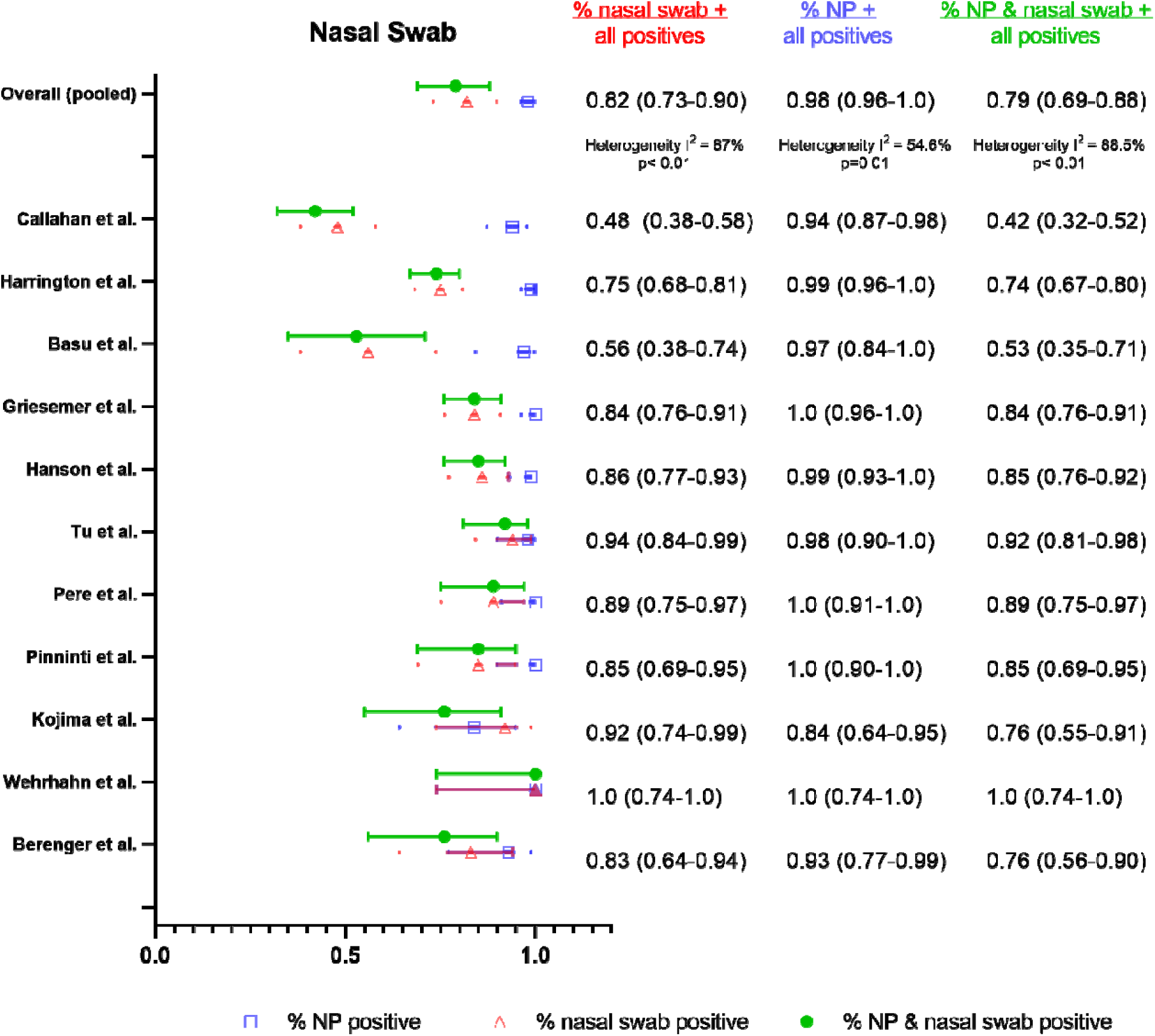
Summary forest plot of individual studies assessing nasal swabs

The pooled estimates of % positive NS via AN (35, 62) versus MT swabs (using the depth of insertion to classify AN vs MT sampling, per CDC description) were similar (54, 59, 63, 66–68) [0.90 (95% CI: 0.84-0.94) vs. 0.84 (95% CI: 0.65 – 0.97), Fig. 5]. % positive NP was similar between groups [0.99 (95% CI: 0.95-1.0) vs. 0.97 (95% CI: 0.92-1.0) respectively]. There were only two studies that assessed AN swabs, and notably, one of the studies (62) also compared AN to MT sampling (AN performed before MT). Performance was similar between AN and MT swabs (48 out of 51 total positives for AN vs. 50 out of 52 total positives for MT). In this study, both the AN and MT swabs were collected from both nares, and the AN swab was unflocked, whereas the MT swab was flocked.

**Fig. 5:**
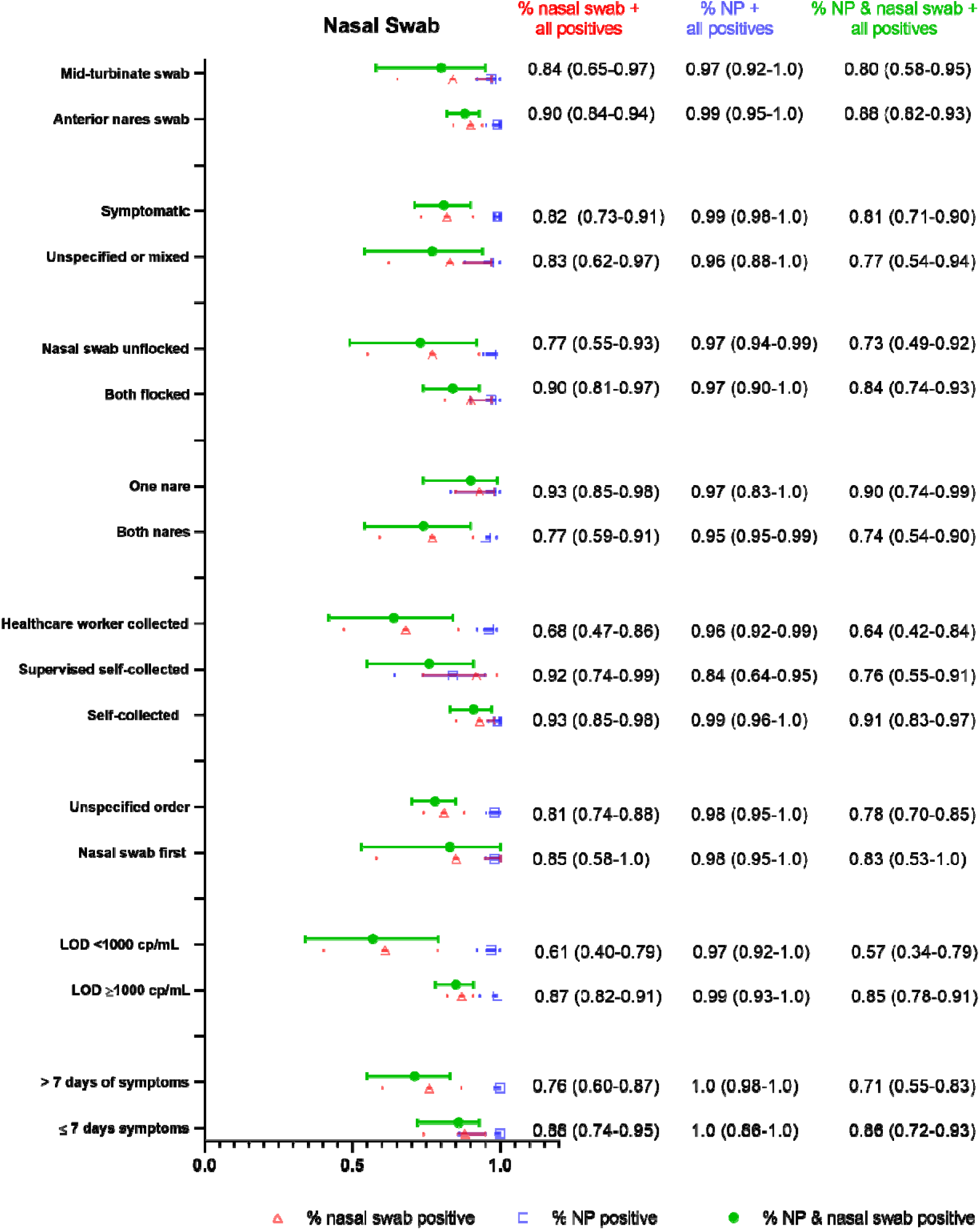
Summary forest plot of sub-group data from nasal swab sampling for different clinical populations, and collection as well as processing procedures.

We found that a more sensitive assay (LOD <1000 copies/mL) (64, 65, 68) resulted in worse performance of NS in comparison to assays with LOD ≥ 1000 copies/mL (32, 54, 59, 63, 66, 67) [0.61 (95% CI: 0.40-0.79) vs. 0.85 (95% CI: 0.82-0.91)], while % positive NP were similar (0.99 (95% CI: 0.93-1.0) vs. 0.97 (95% CI: 0.92-1.0) respectively). This may reflect lower viral burden in the mid-turbinate/anterior nares region than the nasopharynx resulting in lower performance in comparison to NP swabs that is only evident when using a highly sensitive (LOD < 1000 copies/mL) assay. Notably, the discordant paired samples described in the studies also were found to have lower viral loads than concordant pairs.

Two of the studies (64, 68) with the worst NS performance used a more sensitive assay and also compared unflocked NS to flocked NP swabs (positive NS detection 48– 56%). There were 3 other studies (35, 54, 59), however, that also used unflocked NS compared to flocked NP swabs and reported higher detection (83-100%). Ultimately, NS studies that specified that both NS and NP swabs were flocked (63, 66) had a substantially higher % positive NS than studies where an unflocked NS was compared to a flocked NP swab (35, 54, 59, 64, 68), although confidence intervals were wide and overlapping (0.90 (95% CI: 0.81-0.97) vs. 0.77 (95% CI 0.55-0.93), Fig. 5), and this finding may have been partially driven by the two studies using the lower LOD assay. % positive NP between these two groups were similar [0.97 (95% CI: 0.90-1.0) vs. 0.97 (95% CI: 0.94-0.99)]. One study (62) collected unflocked AN swabs, flocked MT swabs, and unflocked NP swabs for comparison and reported similar performance of all three specimens as described above. However, use of a non-flocked swab for the NP sampling may have decreased its sensitivity and artificially increased the sensitivity of MT and AN sampling.

Only 4 studies (35, 59, 62, 68) specified that NS were performed before NP swabs and although % positive NS was higher in comparison to studies that did not specify the swab order, there was not a substantial difference [0.85 (95% CI: 0.58-1.0) vs. 0.81 (95% CI: 0.74-0.88), Fig. 5]. % positive NP was the same regardless of NS order [0.98 (95% CI: 0.95-1.0)]. Surprisingly, NS specimens collected from both nares (35, 54, 62, 64, 67, 68) seemed to perform worse in comparison to swabs collected from a single nostril (59, 63, 66), although the difference was not substantial [0.77 (95% CI: 0.59-0.91) vs. 0.93 (0.59-0.91), Fig. 5]. % positive NP was again similar in these two scenarios [0.95 (95% CI: 0.95-0.99) for both nares vs. 0.97 (95% CI: 0.83-1.0) for one nare]. Notably, this finding may again have been driven by the two studies (64, 68) utilizing a more sensitive < 1000 cp/mL NAAT assay (both with poor detection by NS and performed with swabs collected in both nares), a factor which we described previously as associated with a lower NS detection rate.

We also found that unsupervised self-collected NS specimens (35, 59, 62) had higher percent positives in comparison to swabs collected by healthcare workers (54, 64, 67, 68) [0.93, 95% CI: 0.85-0.98 vs. 0.68, 95% CI: 0.47-0.86, Fig. 5]. In only one study (63), the patient’s self-collection was supervised (0.92, 95% CI: 0.74-0.99). Professional NP sampling performance was similar between the NS self-collection and HCW-collection groups [0.99 (95% CI: 0.96-1.0) vs. 0.96 (95%CI: 0.92-0.99)]. Notably, the same two studies (64, 68) showing the worst NS performance (and using more sensitive assays) were in the healthcare-worker collected group.

We could not find any studies that specifically assessed only asymptomatic patients with NS, although multiple studies used mixed populations and a few studies specified that all of the cohort was symptomatic. Studies of only-symptomatic patients (35, 62, 64–67) had a similar % positive NS rate to studies of mixed or unspecified patients [0.82 (95% CI: 0.73-0.91) vs. 0.83 (95% CI: 0.62-0.97), Fig. 5]. % positive NP was similar between groups [0.99 (95% CI: 0.98-1.0) vs. 0.96 (95% CI: 0.88-1.0)]. There were two studies (63, 67) with extractable data to assess the impact of symptom duration prior to testing, and a lower yield after 7 days was found although the difference was not substantial [0.76 (95% CI: 0.60-0.87) for > 7 days symptoms vs. 0.88 (95% CI: 0.74-0.95) for ≤ 7 days, Fig. 5]. % positive NP was similar in both groups [1.0 (95% CI: 0.86-1.0) vs. 1.0 (0.98-1.0)].

Again, data were limited on pediatric populations when using nasal swabs. There are two pediatric studies of nasal samples; one study (69) described NS to be outperforming OP swabs in 56 paired samples from 11 pediatric patients, with Ct values lower in NS versus OP for all 11 first paired samples. This study had to be excluded due to repeat sampling, as we were not able to extract unique patient data from different timepoints. The other study (70) is not a nasal swab study, but describes NP aspirates to be outperforming NP swabs for detection (% positive for NP aspirates was 88% in comparison to 51% for NP swabs), though methods details provided were minimal and repeat sampling on patients occurred.

### Combined OP/NS as a specimen type

Four studies (59, 71–73) evaluated combined oropharyngeal and nasal swabs in comparison to NP swabs (Fig. 6). Three of these studies (71–73) specified that a single swab was used for collection of an OP/NS sample, whereas in one study it was unclear if two separate swabs were used for OP and NS sampling and the results compiled (59). Two of the studies (59, 73) used MT swab depth, while one used AN sampling (72), and one was unspecified (71). Three of the studies were healthcare-collected swabs, in which the OP sampling was performed prior to NS (71–73), and only two of these studies specified swabbing both nares (72, 73). Two studies used flocked swabs (71, 73), while the others used unflocked swabs. The LOD of the assay in copies/mL was only available in one study (59) and > 1000 cp/mL. Two of the studies (59, 71) found that the percent positive detection of the combined swab specimen was greater than percent positive detection for the reference NP swabs. Pooled detection estimates were similarly high between the combined swabs and NP swabs with the same % positive estimate [0.97 (95% CI: 0.90 – 1.0)] although agreement between the two methods was less [0.90 dual positive (95% CI: 0.84 – 0.96)].

**Fig. 6:**
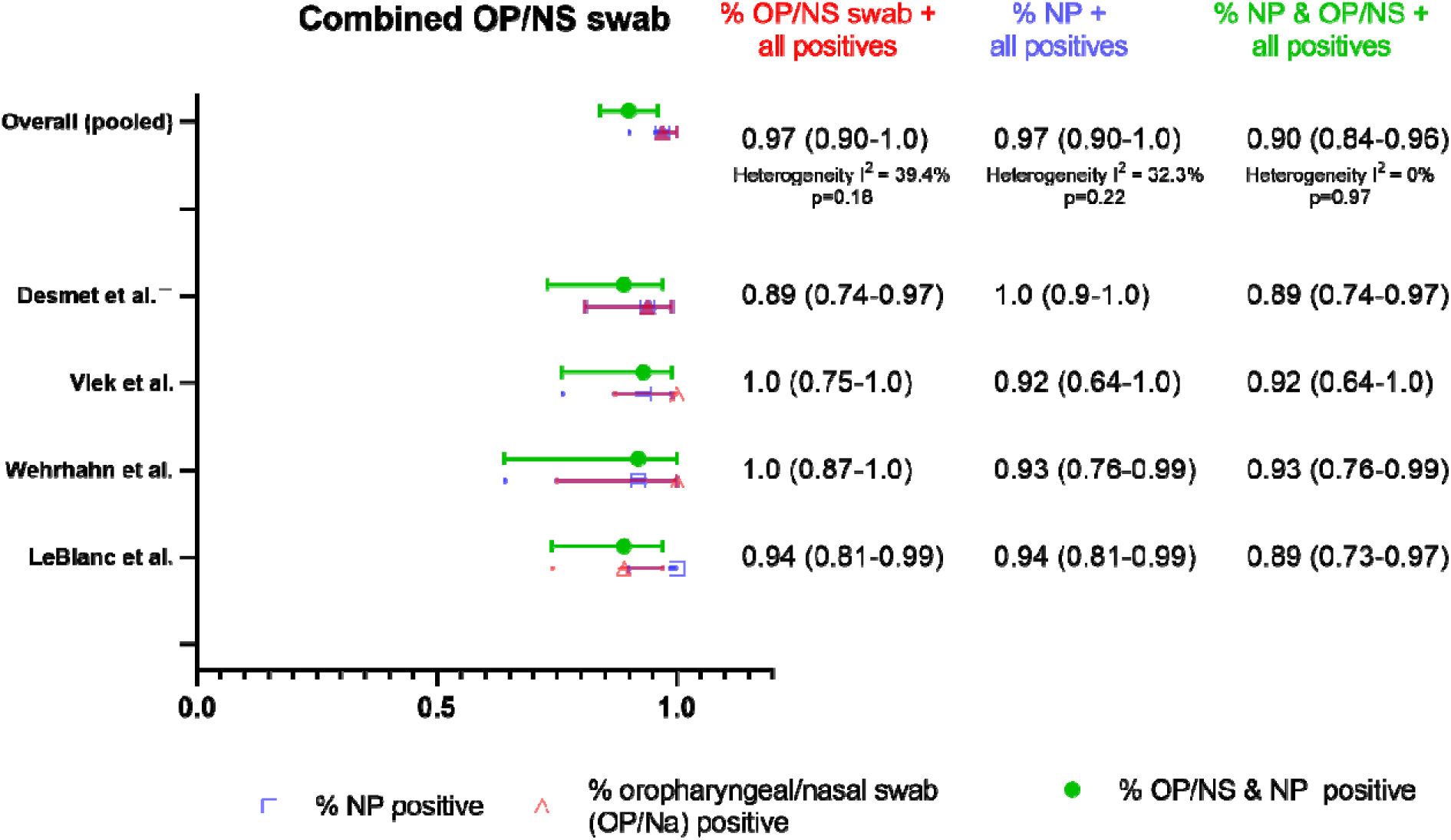
Summary forest plot of individual studies assessing oropharyngeal/nasal swab sampling

### Assessment of publication bias

Visual inspection of a funnel plot of the study data versus standard error shows substantial asymmetry and therefore suggest publication bias for all alternative sample types (Supplementary Fig. 3).

## Discussion

This systematic review and meta-analysis synthesizes a large number of studies comparing alternative sample types to NP swab for SARS-CoV-2 detection by NAAT. While all 3 sample types independently seemed to capture lower % positives [nasal swabs (0.82, 95% CI: 0.73-0.90), OP swabs (0.84, 95% CI: 0.57-1.0) and saliva (0.88, 95% CI: 0.81 – 0.93)] in comparison to NP swabs, combined OP/nasal swabs in 4 studies interestingly had the same % positive detection rate as NP swabs (0.97, 95% CI: 0.90-1.0) (Fig. 6).

For saliva specimens, we found slightly lower performance compared to NP swabs overall [0.88 (95% CI: 0.81-0.93) vs. 0.94 (95% CI: 0.90-0.98)], and only the absence of nucleic acid extraction resulted in a substantially lower rate of detection. One study (44) had to be excluded from the meta-analysis due to non-synchronous collection of NP and saliva samples. This study is of interest nevertheless, as it tested different RT-PCR platforms on the same set of saliva specimens using 3 different extraction-free commercial RT-PCR assays against a standard RT-PCR assay with extraction and reported that 79, 81, and 52 specimens were detected, respectively, out of a total of 84 positive specimens detected on the standard assay, indicating that choice of extraction-free assay matters.

Self-collection, coughing or deep throat saliva, and avoiding food, drink, or toothbrushing resulted in >5% increased positive saliva detection rates, although the difference was not substantial. Collection of saliva greater than 7 days after symptom onset also resulted in >10% lower detection, although the difference was not substantial. Viscosity has been described qualitatively in multiple studies as a challenge in utilization of saliva as a specimen type, but invalid rates were not documented in many studies; we note that invalid rates are a critical parameter and should be consistently reported.

We found that OP swabs seemed to perform similarly to saliva and NP swabs, but these estimates were highly affected by one study (56) where samples were collected near the end of a hospitalization for discharge purposes and the % positive NP was unusually low in comparison to the rest of the literature. Overall, the data argue against self-collection of this sample type.

For NS, the literature to date supports that they perform worse than NP swabs although these findings were largely driven by two studies (64, 68). There was no substantial difference between AN and MT swab detection, and we note that AN/MT swab collection protocols may have overlapped in practice; we defined AN vs MT based on depth of insertion, as the swab type was not always specified. We hypothesize that the difference in NS to NP performance was driven by use of a particularly sensitive (< 1000 cp/mL LOD) assay for two of the studies; while this may reflect differences in viral burden anatomically between NP and AN/MT sampling, the clinical significance of this difference remains to be determined. We note that the discordant samples (NP+/NS-) typically had low viral loads on the NP sample.

We hypothesize that nasal swab performance is likely highly dependent on collection procedure, which has in turn evolved over time and a substantial amount of data remained unpublished (personal communication Nira Pollock). Self-collection and single-nare sampling trended towards higher although not substantially different % positive detection, although these results were largely driven by the two studies with poor NS performance (64, 68). Detection was also lower after > 7 days of symptoms although this difference was not substantial. There remain unresolved questions on the best performing swab material (spun polyester, foam, rayon) and sample transport (buffer, dry swab) that could not be addressed in this study due to limited data (many studies did not report the swab product or elution details). We also still do not fully understand the impact of flocking on this specimen type. While studies using flocked nasal swabs had a slightly higher % positive detection in comparison to unflocked swab studies, the unflocked swab group included two studies using a lower LOD assay, and the 3 studies that used a less sensitive assay had near 90% detection (similar to the flocked group).

OP/NS samples had surprisingly high performance compared to NP swabs; while this sample type likely requires an operator for collection, additional studies are warranted to understand acceptability in adult and pediatric patients.

Overall, much remains unknown about the impact on diagnostic sensitivity of variations in specimen collection and processing protocols, and performance in specific key sub-populations (asymptomatic vs. symptomatic, pediatric vs. adult, late vs. early testing from symptom onset). While we assess aggregate data from different studies to gain insight into these variables, a limitation of this meta-analysis is that true comparison is precluded in the absence of head-to-head studies. Furthermore, while there are trends we observe in our subgroup analyses, these findings may be population-related and should be interpreted with caution. Timing of sampling from symptom onset was also quite variable (collection occurred within days to weeks in some studies), and was inconsistently reported, which likely had a major impact on diagnostic performance given decreasing viral load over time. Head-to-head studies are urgently needed of flocked vs unflocked swabs (and specialized vs unspecialized swabs for MT collection), collected at different times in disease and with different sampling methods, and also in important subpopulations (e.g. children), to resolve the persistent uncertainty.

We note that the reporting quality of studies was low, STARD guidelines (74) were not consistently followed, and study bias was considered moderate to high on QUADAS 2. Lastly, in this study we chose to report the % positive alternative-specimen, % positive comparator-specimen, and % dual positives instead of the positive percent agreement (PPA). This decision was motivated by our presumption regarding the low rate of false-positives using NAAT, and the potential for an alternative to yield more positive results than the comparator NP, which would otherwise not be considered.

In summary, while alternative specimens (particularly saliva and OP/NS samples) show promise, we find that the literature to date suggests that NP swabs are indeed the gold standard in comparison to alternative specimen types (saliva, OP swab, NS). We identify self-collected nasal swabs and saliva to have similar performance to healthcare-worker obtained specimens, which is also supported by a head-to-head comparison of self-collected AN versus professionally-collected NP swabs utilizing an antigen rapid test for detection (75). We reiterate that the LOD of any assay will impact detection and centers should be aware of the increased possibility of false negatives with any sample type when using a less sensitive assay. Given the promising results of combined oropharyngeal and nasal swab studies, more studies on alternative specimen combinations would be useful. Lastly, we encourage future studies to provide more clarity about exact details of collection procedures, specific swab shape and materials used, sample processing methods (dilution, extraction, storage, transport), and NAAT assay utilized (including LOD), allowing the field to clearly define the tradeoffs required to sufficiently bring SARS-CoV-2 testing to scale.

## Data Availability

All data are available upon request and are publicly available from published studies.

## Supplementary Methods

### Example search method employed in systematic review and meta-analysis

The following strategy was used in Medline/Pubmed to identify articles providing a quantitative evaluation of diagnostic tests by specimen type: *(“COVID-19 diagnostic testing”[MeSH Supplementary Concept]* AND “Coronavirus Infection” [MeSH Major Topic] AND [“saliva”[MeSH Major Topic] OR “nose” [MeSH Major Topic] OR “nasal” “oropharynx” [MeSH Major Topic] OR “oropharyngeal” OR “oral” OR “nasopharynx” [MeSH Major Topic] OR “nasopharyngeal”]. We also searched the grey literature via Google Scholar as well as Medrvix and bioRxiv via search terms that included a combination of subject headings (when applicable) and text-words for the concepts: (1) Sample type (“saliva” OR “oral” OR “oropharyngeal” OR “nasopharyngeal” OR “nasal” OR “swab”); (2) Diagnosis and (3) Disease (“SARS-COV-2” OR “COVID”).

**Supplementary Table 1:**
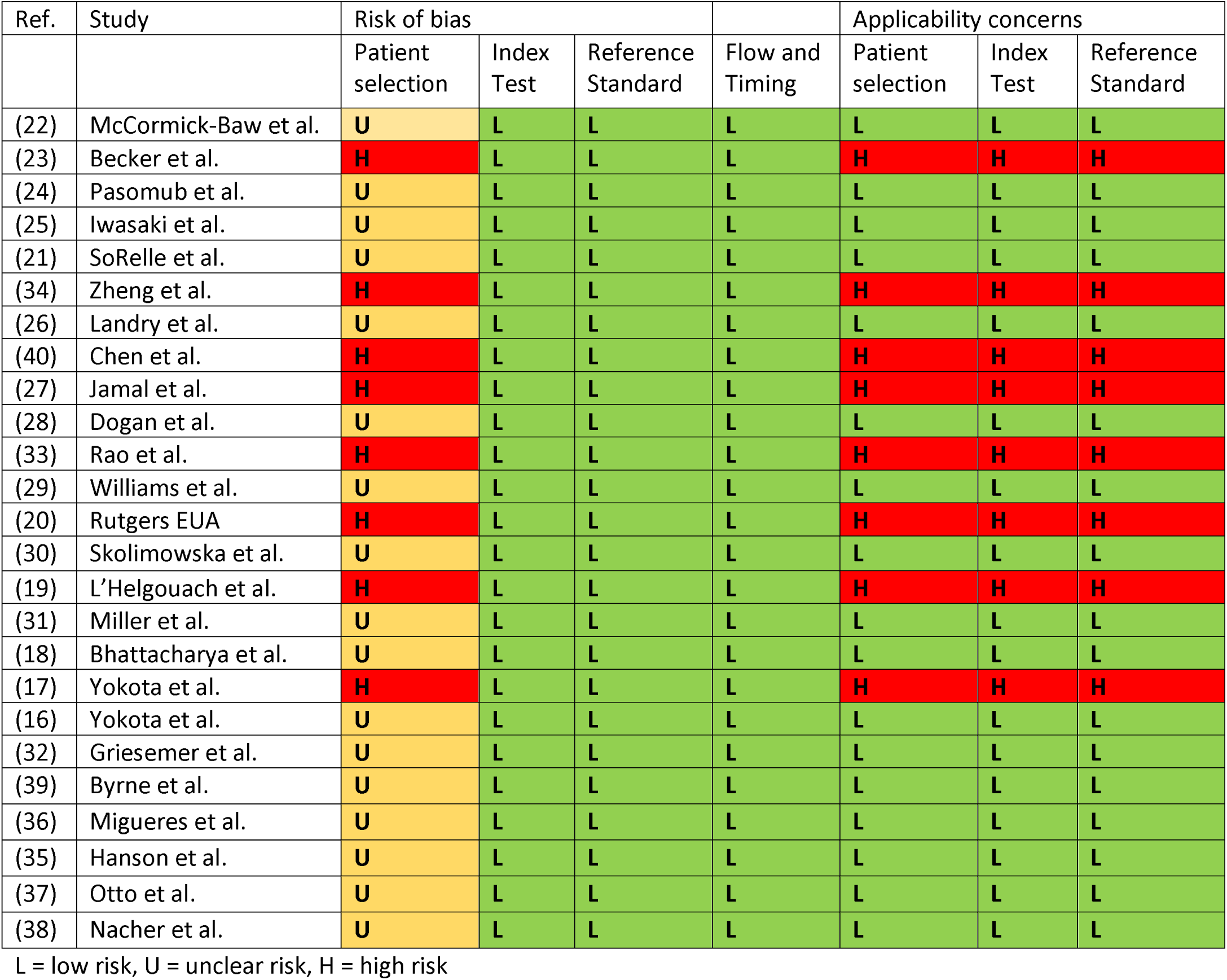
Risk of bias in saliva studies

**Supplementary Table 2:** Risk of bias in OP swab studies

| Ref. | Study | Risk of bias |  |  |  | Applicability concerns |  |  |
| --- | --- | --- | --- | --- | --- | --- | --- | --- |
|  |  | Patient selection | Index Test | Reference Standard | Flow and Timing | Patient selection | Index Test | Reference Standard |
| Oral swabs |  |  |  |  |  |  |  |  |
| (54) | Berenger et al. | H | L | L | L | H | L | L |
| (59) | Wehrhahn et al. | U | L | L | L | L | L | L |
| (55) | Wang et al. | H | L | L | L | H | L | L |
| (56) | Yu et al. | H | L | L | L | H | L | L |
| (57) | Calame et al. | H | L | L | L | H | L | L |
| (58) | Patel et al. | U | L | L | U | L | L | L |

L = low risk, U = unclear risk, H = high risk

**Supplementary Table 3:** Risk of bias in nasal swab studies

| Ref. | Study | Risk of bias |  |  |  | Applicability concerns |  |  |
| --- | --- | --- | --- | --- | --- | --- | --- | --- |
|  |  | Patient selection | Index Test | Reference Standard | Flow and Timing | Patient selection | Index Test | Reference Standard |
| (54) | Berenger et al. | U | L | L | L | L | L | L |
| (59) | Wehrhahn et al. | U | L | L | H | L | L | L |
| (63) | Kojima et al. | U | L | L | L | H | L | L |
| (67) | Pinninti et al. | H | L | L | L | H | L | L |
| (66) | Pere et al. | U | L | L | L | L | L | L |
| (62) | Tu et al. | U | L | L | L | L | L | L |
| (35) | Hanson et al. | U | L | L | L | L | L | L |
| (32) | Griesemer et al. | U | L | L | L | L | L | L |
| (64) | Basu et al. | U | L | L | L | L | L | L |
| (65) | Harrington et al. | L | L | L | L | L | L | L |
| (68) | Callahan et al. | U | L | L | L | L | L | L |
| Combined oropharyngeal and nasal swabs |  |  |  |  |  |  |  |  |
| (72) | LeBlanc et al. | U | L | L | U | L | L | L |
| (59) | Wehrhahn et al. | U | L | L | L | L | L | L |
| (71) | Vlek et al. | U | L | L | L | L | L | L |
| (73) | Desmet et al. | U | L | L | L | L | L | L |

L = low risk, U = unclear risk, H = high risk

**Supplementary Table 4:** Saliva collection procedures specified in methodology for meta-analysis studies

|  | n/25 total studies (%) |
| --- | --- |
| Coughing before collection<br>(34, 37, 40) | 3 (12%) |
| Drooling or spitting<br>(16–32, 35, 36, 38, 39, 51) | 22 (88%) |
| Specified deep throat or posterior oropharyngeal (33) | 2 (8%) |
| Avoiding food, drink, brushing teeth<br>(22, 23, 26, 31–34, 40) | 10 (40%) |
| Morning submission (33, 40) | 2 (8%) |
| Nucleic acid extraction free<br>(19, 28) | 2 (8%) |
| Diluted<br>(16, 17, 23–25, 27–32, 35, 37, 40) | 14 (56%) |
| Undiluted<br>(22, 26) | 2 (8%) |
| Assay LOD < 1000 copies/mL<br>(20, 21, 24, 34, 39) | 5 (20%) |
| Assay LOD ≥ 1000 copies/mL<br>(23, 26, 27, 31–33, 51) | 7 (28%) |
| Self-collection<br>(16, 17, 20, 25–27, 29, 30, 33, 34, 37, 39, 40) | 13 (52%) |
| Supervised or HCW collected saliva<br>(22, 28, 31, 35, 38) | 5 (20%) |
| Asymptomatic patients<br>(16, 19, 33) | 6 (24%) |
| Symptomatic patients<br>(17, 18, 21–23, 25, 27, 28, 34, 35, 37, 39, 40) | 13 (52%) |

**Supplementary Table 5:** Nasal swab collection procedures specified in methodology for meta-analysis studies

|  | n/11 total studies (%) |
| --- | --- |
| Nasal swab first before NP swab<br>(35, 59, 62, 68) | 4 (36%) |
| Anterior nares swab<br>(35, 62) | 10 (91%) |
| Mid-turbinate nares swabs<br>(54, 59, 63, 66–68) | 6 (55%) |
| Both swabs flocked<br>(63, 66) | 2 (18%) |
| Nasal swab unflocked in comparison to flocked NP swab<br>(35, 54, 59, 64, 68) | 5 (45%) |
| Both nares<br>(35, 54, 62, 64, 67, 68) | 6 (56%) |
| One nare<br>(59, 63, 66) | 3 (27%) |
| Assay LOD < 1000 copies/mL<br>(64, 65, 68) | 3 (27%) |
| Assay LOD ≥ 1000 copies/mL<br>(32, 54, 59, 63, 66, 67) | 6 (55%) |
| Self-collected nasal swab<br>(35, 59, 62) | 3 (27%) |
| Supervised nasal swab collection (63) | 1 (9.1%) |
| HCW collected saliva<br>(54, 64, 67, 68) | 4 (36%) |
| Symptomatic patients<br>(35, 62, 64–67) | 6 (55%) |

**Supplementary Fig. 1:**
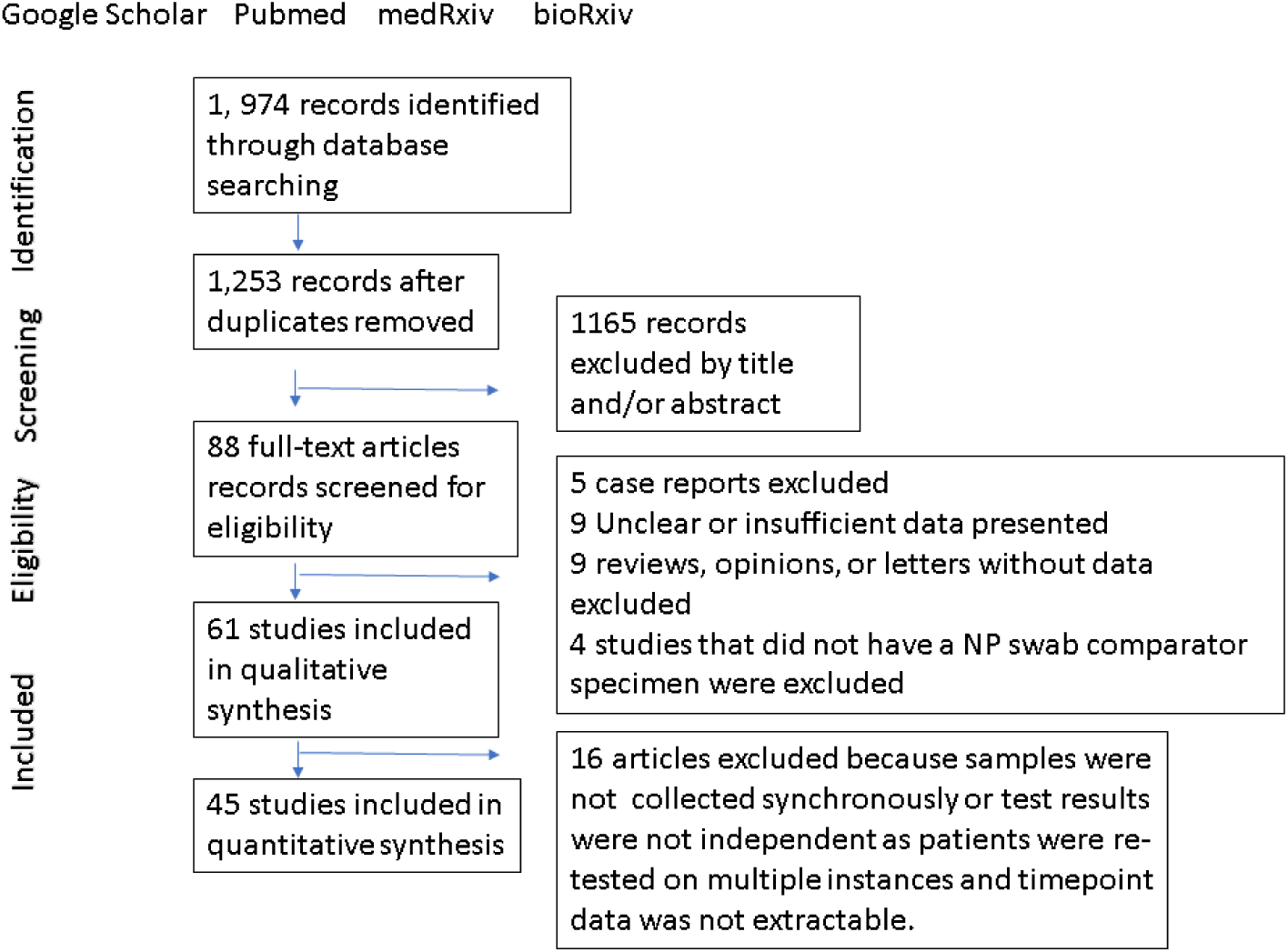
Study retrieval diagram

**Supplementary Fig. 2:**
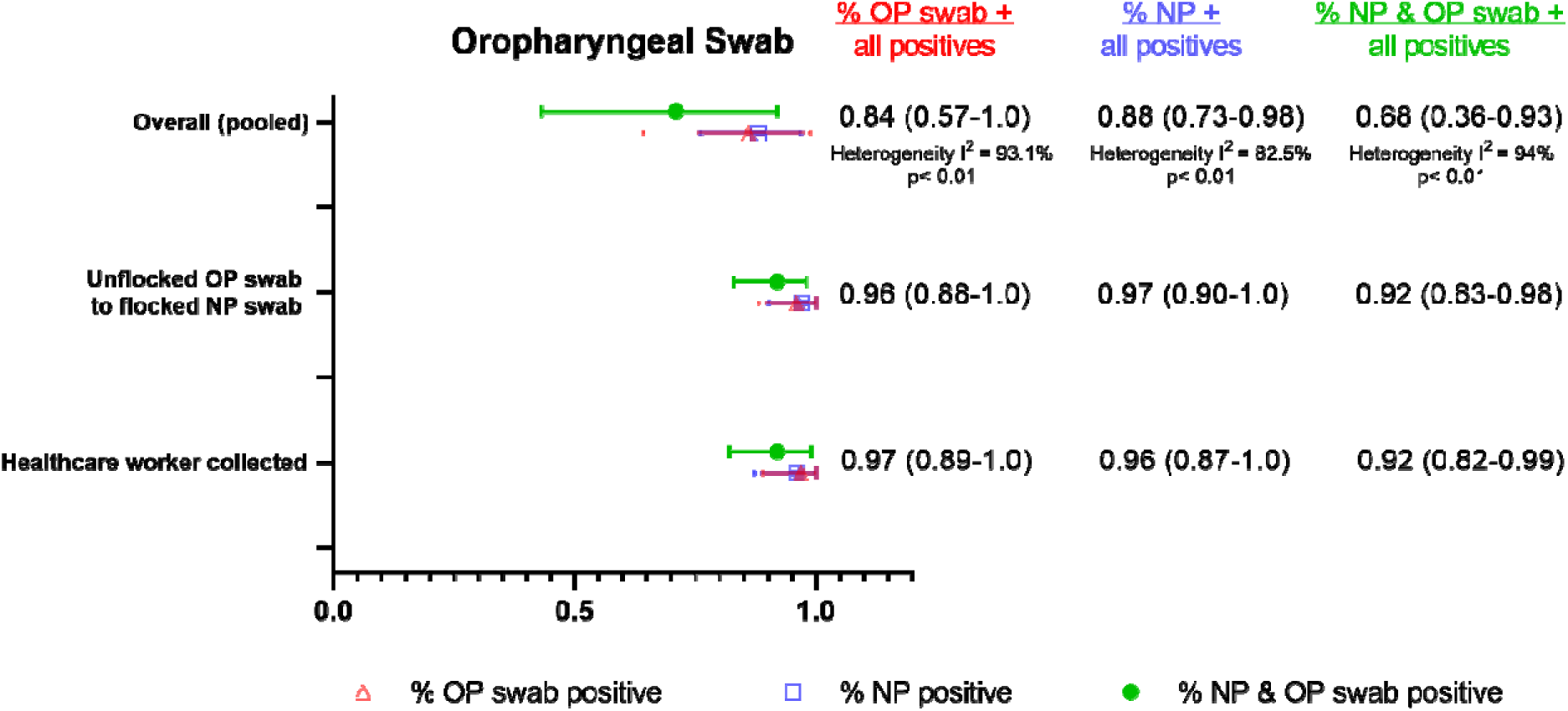
Summary forest plot of sub-group data from OP swabs

**Supplementary Fig. 3:**
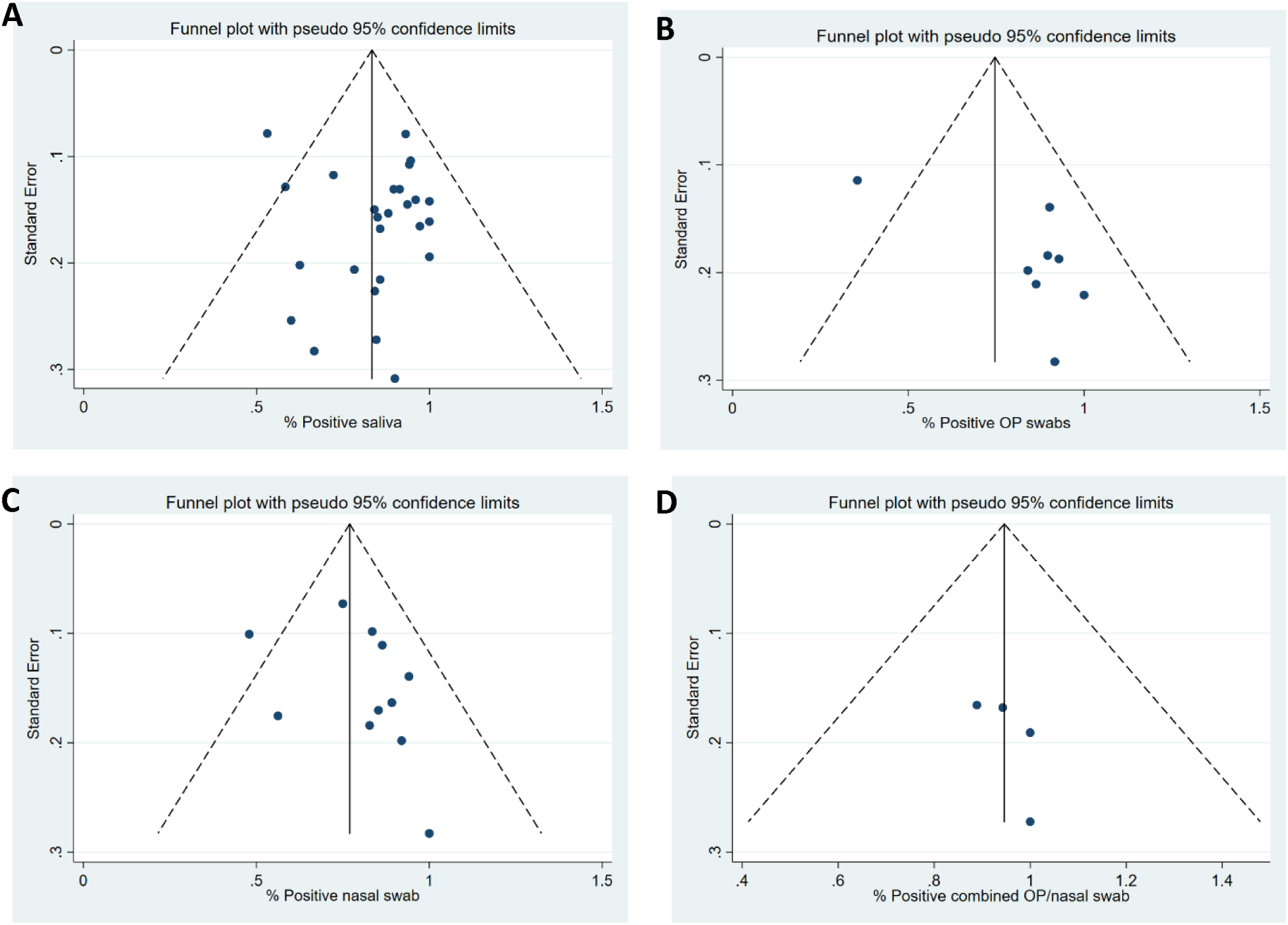
Funnel plots for saliva, OP, nasal, and OP/nasal swabs studies respectively (A-D).

Supplementary File: PRISMA Checklist

| Section/topic | # | Checklist item | Reported on page # |
| --- | --- | --- | --- |
| **TITLE** |  |  |  |
| Title | 1 | Identify the report as a systematic review, meta-analysis, or both. | 1 |
| **ABSTRACT** |  |  |  |
| Structured summary | 2 | Provide a structured summary including, as applicable: background; objectives; data sources; study eligibility criteria, participants, and interventions; study appraisal and synthesis methods; results; limitations; conclusions and implications of key findings; systematic review registration number. | 2 |
| **INTRODUCTION** |  |  |  |
| Rationale | 3 | Describe the rationale for the review in the context of what is already known. | 3 |
| Objectives | 4 | Provide an explicit statement of questions being addressed with reference to participants, interventions, comparisons, outcomes, and study design (PICOS). | 4 |
| **METHODS** |  |  |  |
| Protocol and registration | 5 | Indicate if a review protocol exists, if and where it can be accessed (e.g., Web address), and, if available, provide registration information including registration number. | 4 |
| Eligibility criteria | 6 | Specify study characteristics (e.g., PICOS, length of follow-up) and report characteristics (e.g., years considered, language, publication status) used as criteria for eligibility, giving rationale. | 4 |
| Information sources | 7 | Describe all information sources (e.g., databases with dates of coverage, contact with study authors to identify additional studies) in the search and date last searched. | 4 |
| Search | 8 | Present full electronic search strategy for at least one database, including any limits used, such that it could be repeated. | Suppl, Methods |
| Study selection | 9 | State the process for selecting studies (i.e., screening, eligibility, included in systematic review, and, if applicable, included in the meta-analysis). | 4 |
| Data collection process | 10 | Describe method of data extraction from reports (e.g., piloted forms, independently, in duplicate) and any processes for obtaining and confirming data from investigators. | 4-5 |
| Data items | 11 | List and define all variables for which data were sought (e.g., PICOS, funding sources) and any assumptions and simplifications made. | 4-5 |
| Risk of bias in individual studies | 12 | Describe methods used for assessing risk of bias of individual studies (including specification of whether this was done at the study or outcome level), and how this information is to be used in any data synthesis. | 5 |
| Summary measures | 13 | State the principal summary measures (e.g., risk ratio, difference in means). | 5 |
| Synthesis of results | 14 | Describe the methods of handling data and combining results of studies, if done, including measures of consistency (e.g., $I^2$) for each meta-analysis. | 5 |
| Risk of bias across studies | 15 | Specify any assessment of risk of bias that may affect the cumulative evidence (e.g., publication bias, selective reporting within studies). | 5 |
| Additional analyses | 16 | Describe methods of additional analyses (e.g., sensitivity or subgroup analyses, meta-regression), if done, indicating which were pre-specified. | 5 |
| **RESULTS** |  |  |  |
| Study selection | 17 | Give numbers of studies screened, assessed for eligibility, and included in the review, with reasons for exclusions at each stage, ideally with a flow diagram. | 6 |
| Study characteristics | 18 | For each study, present characteristics for which data were extracted (e.g., study size, PICOS, follow-up period) and provide the citations. | 6-11 |
| Risk of bias within studies | 19 | Present data on risk of bias of each study and, if available, any outcome level assessment (see item 12). | 11, Suppl Tables 1-3 |
| Results of individual studies | 20 | For all outcomes considered (benefits or harms), present, for each study: (a) simple summary data for each intervention group (b) effect estimates and confidence intervals, ideally with a forest plot. | 6-11, Fig. 1-6 |
| Synthesis of results | 21 | Present results of each meta-analysis done, including confidence intervals and measures of consistency. | 6-11, Fig.1-6 |
| Risk of bias across studies | 22 | Present results of any assessment of risk of bias across studies (see Item 15). | Supp. Fig. 3 |
| Additional analysis | 23 | Give results of additional analyses, if done (e.g., sensitivity or subgroup analyses, meta-regression [see Item 16]). | 6-11 |
| **DISCUSSION** |  |  |  |
| Summary of evidence | 24 | Summarize the main findings including the strength of evidence for each main outcome; consider their relevance to key groups (e.g., healthcare providers, users, and policy makers). | 11-13 |
| Limitations | 25 | Discuss limitations at study and outcome level (e.g., risk of bias), and at review-level (e.g., incomplete retrieval of identified research, reporting bias). | 12 |
| Conclusions | 26 | Provide a general interpretation of the results in the context of other evidence, and implications for future research. | 13 |
| **FUNDING** |  |  |  |
| Funding | 27 | Describe sources of funding for the systematic review and other support (e.g., supply of data); role of funders for the systematic review. | 13 |

